# Parietal Default Mode Network Connectivity is Associated with Tobacco Use in Psychosis

**DOI:** 10.64898/2026.03.02.26347415

**Authors:** Yunong Bai, Andrew R. Kittleson, Baxter P. Rogers, Anna Huang, Neil D. Woodward, Stephan Heckers, Julia M. Sheffield, Simon Vandekar, Heather Burrell Ward

**Author notes:** <u>Corresponding Author:</u> Heather Burrell Ward, MD, 1601 23^rd^ Ave S, Nashville, TN 37212.

## Abstract

**Background and Hypothesis:** Abnormal default mode network (DMN) connectivity was observed in both tobacco use and psychotic spectrum disorders, but it remains unknown how psychosis impacts the relationship between connectivity and tobacco use. Interventions targeting the left lateral parietal DMN node (LLP_DMN_) have modulated DMN connectivity and nicotine craving in psychosis. We aimed to investigate relationships between DMN connectivity, psychotic illness, and tobacco use.

**Study Design:** 336 participants (psychosis: n=161, control: n=175) reported their tobacco use history and underwent resting-state functional magnetic resonance imaging. We calculated connectivity within DMN and salience network (SN), between DMN-SN, and from LLP_DMN_ to other DMN and SN nodes. Logistic and LASSO regression with bootstrapping were performed to investigate diagnosis-by-connectivity interactions on lifetime tobacco use. Exploratory brainwide analysis was conducted by regressing brainwide connectivity to LLP_DMN_ against daily cigarette use.

**Study Results:** We observed a significant diagnosis-by-DMN connectivity interaction for lifetime tobacco use (p=0.0281, coefficient=0.457, OR=1.579, 95% CI=[1.063, 2.411]); in the psychosis group, higher DMN connectivity was associated with higher odds of lifetime tobacco use. LASSO regression yielded four predictors of lifetime tobacco use: age, diagnosis, LLP_DMN_ connectivity to a prefrontal SN node, and the interaction between diagnosis and LLP_DMN_ connectivity to a right parietal DMN node. Brainwide analysis identified bilateral somatomotor clusters where higher connectivity to LLP_DMN_ correlated with higher daily cigarette use (voxel-wise p<0.001, cluster p<0.05).

**Conclusions:** Psychosis diagnosis modified relationship between DMN connectivity and tobacco use. Modulating DMN connectivity may provide a psychosis-specific treatment target for tobacco dependence.

## Introduction

Tobacco use is the leading preventable cause of early mortality in psychosis spectrum disorders^1^, leading to a 20-year decreased life expectancy in schizophrenia (SZ).^2^ Compared to the general population, individuals with psychotic disorders have a higher prevalence of tobacco use^3,4^ and more severe nicotine dependence^5^. However, current smoking cessation interventions are derived from non-psychosis populations, despite evidence that both pharmacologic and neuromodulation treatments for smoking cessation are less effective in psychotic disorders.^6^ There remains a significant gap in our understanding of the neurobiological basis and treatment of tobacco dependence in psychotic disorders, and more effective treatments are desperately needed.

Functional neuroimaging has been used to study the neurobiological mechanisms of tobacco dependence in psychosis but has not converged on an underlying, unified basis for the high prevalence of tobacco use in psychotic disorders. On the network level, much of the existing research has focused on abnormalities in the reward circuitry in psychosis. However, an agnostic, data-driven analysis observed a novel link between tobacco dependence and altered organization of the default mode network (DMN), a resting-state brain network involved in self-referential thoughts^7^ and linked to cognitive performance.^8^ This relationship was specific to SZ and not observed in smokers without a psychotic disorder,^9^ suggesting a potential avenue for studying psychosis-specific mechanisms of and interventions for tobacco dependence.

Aberrant DMN organization has been observed in both psychotic disorders and in tobacco use. Individuals with SZ have higher DMN connectivity at rest,^10^ reduced suppression of DMN activity during cognitive tasks,^8^ and weaker connectivity between DMN and salience network (SN).^11^ Abnormal DMN connectivity and activation are also observed in schizoaffective disorders^12^ and bipolar disorders.^13,14^ Similarly, tobacco use has been linked to the DMN. In psychosis, higher DMN connectivity was linked to stronger tobacco craving,^15^ and nicotine administration normalized baseline DMN hyperconnectivity.^9^ Decreased DMN connectivity^16^ and altered DMN-SN coactivation have also been reported in non-psychosis individuals.^17,18^ Overall, modulating DMN connectivity may offer a novel and effective treatment target to reduce tobacco use in people with psychotic disorders.

One method to modulate DMN connectivity is using transcranial magnetic stimulation (TMS), a form of noninvasive neuromodulation. Whereas TMS applied to the left dorsolateral prefrontal cortex (DLPFC) is FDA-cleared for smoking cessation in adults without psychosis,^19^ it is less effective in SZ.^20^ Conversely, DMN-targeted TMS applied to the left lateral parietal node of DMN (LLP_DMN_) successfully modulated both DMN connectivity and nicotine craving in SZ,^21^ but not in smokers without psychosis.^22^

However, current studies have yet to establish DMN connectivity as a theoretical basis for tobacco dependence in psychotic disorders. Previous studies investigating DMN connectivity in tobacco use and psychotic disorders are few, have small sample sizes, not widely replicated, and underpowered to identify a diagnosis interactive effect. Moreover, previous studies are often limited to SZ. Finally, despite evidence for LLP_DMN_-targeted TMS reducing nicotine craving in psychosis, there is limited study characterizing LLP_DMN_ connectivity in psychosis and tobacco use.

Using a large dataset of individuals with psychosis spectrum disorders (i.e., nonaffective and affective psychoses) and non-clinical controls, we aimed to investigate the relationship between DMN connectivity and tobacco use in people with psychosis. Specifically, we conducted hypothesis-driven analyses of network-level (within- and between-DMN/SN) and edge-level (LLP_DMN_ to other DMN/SN nodes) connectivity to examine how psychosis and tobacco use relates to connectivity. We hypothesized that psychosis diagnosis would influence the relationship between connectivity and tobacco use, such that higher DMN connectivity would be associated with more tobacco use in people with psychosis but not in non-clinical controls. We then performed a brainwide exploratory analysis to find regions where connectivity to LLP_DMN_ was associated with daily cigarette use. This study will further our understanding on the neural mechanisms that predispose people with psychosis to tobacco dependence and will also serve as a theoretical basis for a novel DMN-targeted TMS protocol to treat tobacco dependence in people with psychosis spectrum disorders.

## Methods

### Participants

Data came from a repository of 336 individuals with psychotic disorders and non-clinical controls with complete neuroimaging and behavioral data who participated in one of four neuroimaging projects conducted at Vanderbilt University Medical Center (VUMC, CT00762866; 1R01MH070560; 1R01MH102266; VR71021). All studies were approved by the Vanderbilt Institutional Review Board, and all individuals provided written informed consent prior to participating in the studies. See Supplement for Exclusion criteria.

### Assessments

Tobacco use data was collected for all participants, including history of past or current use of smoking tobacco (e.g., cigarettes, bowls, chew, snuff) and current number of cigarettes smoked per day. Participants with lifetime tobacco use were defined as those with past or current use of smoking tobacco, and the rest were defined as with never-use of tobacco. Data on noncombustible nicotine use, such as vaping, was not collected. Psychosis symptoms were measured using the Positive and Negative Syndrome Scale (PANSS)^23^. To investigate differences between early and chronic psychosis, we defined early psychosis as illness duration less than two years.

### MRI Acquisition

Neuroimaging data were collected on one of two identical 3.0-T Philips Intera Achieva MRI scanners (Philips Healthcare, Andover, MA). Briefly, a seven or ten-minute echo-planar imaging resting-state fMRI scan and T1-weighted anatomical (1 mm isotropic resolution) were collected for each participant. For a subset of individuals (n=28), a cognitive task preceded the resting-state scan; see Supplement for task details. For this reason, scan sequence was included as a covariate in subsequent analyses. See Supplement for full scanning parameters.

### MRI Data Processing

Anatomical images were segmented into gray matter, white matter and cerebrospinal fluid (CSF) with the Computational Anatomy Toolbox 12 (CAT12, version 12.5; http://www.neuro.uni-jena.de/cat/). Resting-state scans were preprocessed in SPM12 and were (1) realigned to a mean scan, (2) coregistered with the native space structural scan, (3) spatially normalized to MNI-space using the parameters obtained from the grey matter segmentation normalization. Then, all scans (4) underwent resting-state denoising procedures: bandpass filter (0.01–0.1 Hz), regression of CSF, gray matter, and white matter signal, regression of 12 motion parameters (six translation and rotation parameters and their first derivatives). All resting-state scans went through a quality assurance procedure that included calculating framewise displacement (FD) and temporal signal-to-noise ratio (tSNR). Scans with a mean FD > 0.5 (n=29) or a tSNR lower than the 5th percentile of the distribution of the entire sample (n=20) were excluded from further analysis.^24^ After quality control, there were a total of 336 scans for analysis.

### Calculation of Whole-Network Connectivity Values

We calculated the average connectivity of resting-state networks as previously described.^9^ To calculate individual values of average DMN functional connectivity, we placed 6mm diameter spheres at coordinates corresponding to nine nodes of the DMN as defined by Raichle et al (posterior cingulate/precuneus [PCC], medial prefrontal, left lateral parietal [LLP_DMN_], right lateral parietal [RLP_DMN_], left inferior temporal, and right inferior temporal cortices; medial dorsal thalamus; right posterior cerebellum; and left posterior cerebellum)^25^. The timecourses of the blood-oxygen-level-dependent (BOLD) signal from the regions of interest (ROIs) were z-transformed and correlated with each other to generate a 9×9 ROI-ROI connectivity matrix. Connectivity from each ROI to every other ROI was calculated and averaged, which produced a mean network connectivity value for each participant (excluding the diagonal). We repeated this process using the SN as defined by Raichle et al (dorsal anterior cingulate, left anterior prefrontal [LaPFC], right anterior prefrontal, left insula, right insula, left lateral parietal [LLP_SN_], right lateral parietal [SN RLP_SN_] cortices)^25^ to calculate connectivity within and between networks (all 6mm diameter spheres). See Supplement for MNI coordinates for each node.

### Network- and Edge-Level Analyses

Logistic regression was used to predict tobacco use status (lifetime or never-use) based on the interaction between network-level connectivity (within DMN, within SN, and between-DMN-SN) and diagnosis (psychosis or control), controlling for age, sex, antipsychotic medication dosage (chlorpromazine equivalence), scanner, and scanning protocol. Logistic least absolute shrinkage and selection operator (LASSO) regression was used to predict tobacco use status based on the interaction between edge-level connectivity (from LLP_DMN_ to other DMN and SN nodes) and diagnosis. The dataset was split into training (70%) and testing (30%) sets. A LASSO regression model was fitted on the training set with 10-fold cross-validation. Model performance was evaluated on the held-out testing set using area under the curve (AUC) and sensitivity and specificity of the optimal threshold. To quantify the uncertainty of the model estimates, we used bootstrapping with 1000 replications and 10-fold cross-validation. Bootstrapping calculated the average estimated regression coefficient (beta) selection frequency and confidence intervals for each predictor across all bootstrap replications.

### Unrestricted, Brain-wide Functional Connectivity Analysis

Brainwide analysis was performed on participants with lifetime tobacco use. We calculated voxel-wise connectivity by extracting the time course of the BOLD signal from a 6mm sphere placed at left lateral parietal LLP_DMN_ coordinates (MNI [−46, −66, +30])^25^. Using SPM12 (Statistical Parametric Mapping, http://www.fil.ion.ucl.ac.uk/spm), we regressed the z-transformed Pearson’s correlation coefficient connectivity maps against daily cigarette use, while controlling for age, sex assigned at birth, scanner, and scanning protocol as covariates, to generate spatial maps of how connectivity from the LLP_DMN_ region to the entire brain varied with daily cigarette use at an uncorrected voxelwise threshold of p<0.001 and cluster-forming threshold of p<0.05. Daily cigarette use in participants with past use was designated to be zero.

We used “cluster” to refer to each group of voxels with significant correlation in the generated spatial map, and “peak voxel” to refer to the voxel with the strongest correlation in each cluster. To visualize the relationship between LLP_DMN_–cluster connectivity and daily cigarette use, we plotted region-to-seed connectivity by measuring BOLD correlation between the LLP_DMN_ region and 6mm spheres (seeds) placed at the location of maximal connectivity-cigarette use association in the cluster using Matlab (MathWorks, Inc.).^26^ In a post-hoc analysis, we used linear regression to investigate whether diagnosis modulated the relationship between LLP_DMN_–peak voxel connectivity and daily cigarette use by regressing diagnosis x smoking interaction against connectivity, controlling for age, sex, scanner, and scanning protocol.

We observed that one cluster partially overlapped with the right insula. In another post-hoc analysis, we extracted region-to-seed connectivity by measuring BOLD correlation between LLP_DMN_ and 6mm seeds placed at the right dorsal granular insula using the ROI defined by Fan et al. (MNI [38, −8, 8]).^27^ We used Pearson’s correlation to investigate the relationship between LLP_DMN_–insula connectivity and daily cigarette use, and linear regression to investigate whether diagnosis modulated this relationship, controlling for age, sex, scanner, and scanning protocol.

### Other Statistical Analyses

Chi-squared tests were used to compare differences in categorical variables between psychosis and control participant groups, including race (white, black or African American, or other), sex assigned at birth (male or female) and tobacco use history (current or non-current, ever or never). See Supplement for analyses of group differences in network connectivity by diagnosis, tobacco use, and psychotic illness duration. See supplement for analysis of group differences in psychosis symptom severity by tobacco use. All analyses described above were conducted in RStudio (Version 2025.05.1+513)^28^.

## Results

### People with Psychosis Have a Higher Lifetime Prevalence of Nicotine Use

Sample demographics are summarized in Table 1. 92 participants endorsed current tobacco use (psychosis: n = 75; control: n = 17), 45 endorsed past use (psychosis: n = 31; control: n = 14), and 199 had never used (psychosis: n = 69; control: n = 130). There were no significant differences between the psychosis and control groups in age, sex assigned at birth, or race (all p > 0.1). The psychosis group had a higher proportion of participants with current- and ever-use of tobacco (both p < 0.001). Lifetime tobacco use was not associated with psychotic symptoms (Supplement). DMN, SN, and DMN-SN network connectivity was not associated with diagnosis or tobacco use independently (Supplement).

**Table 1.**
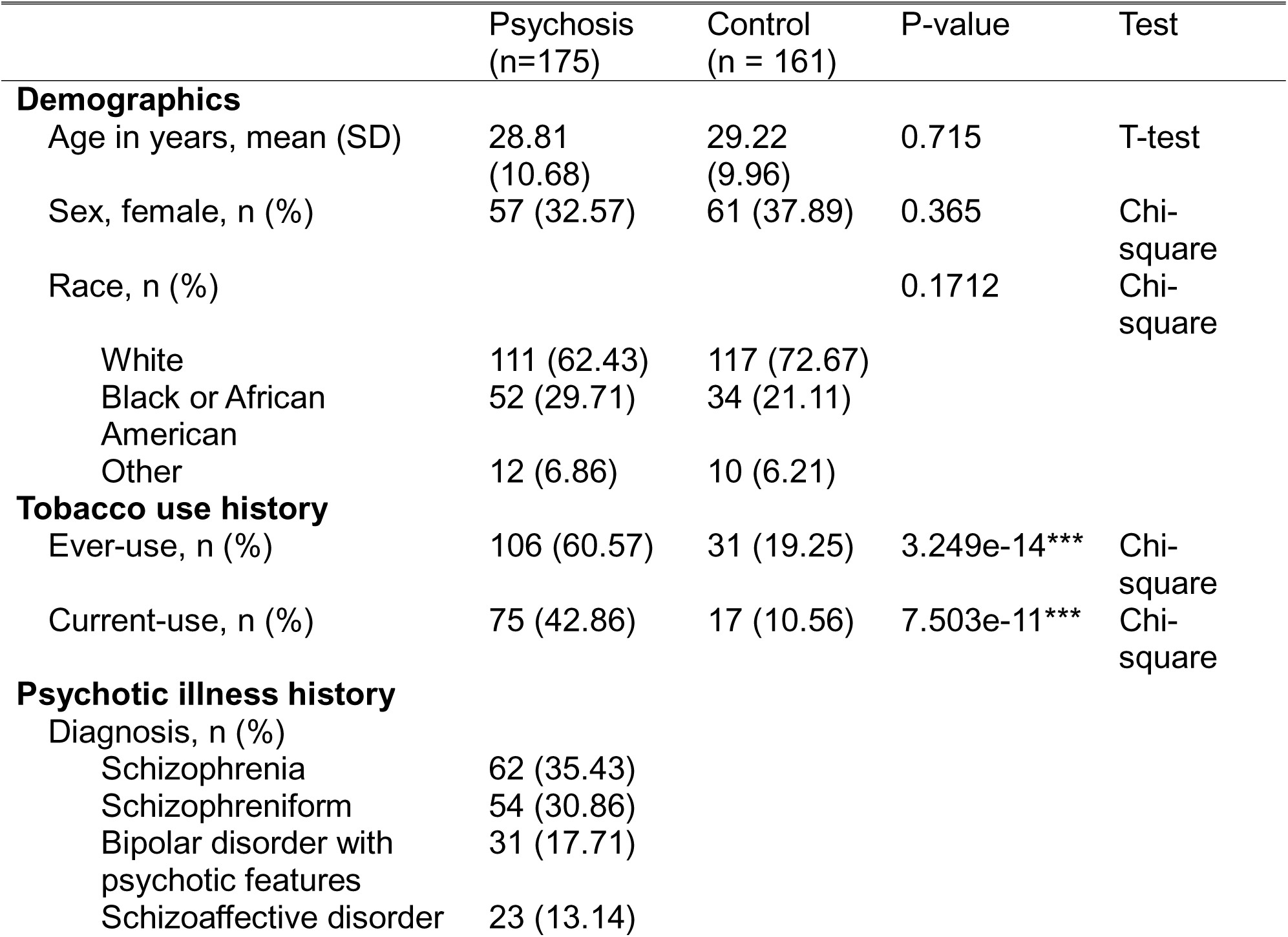
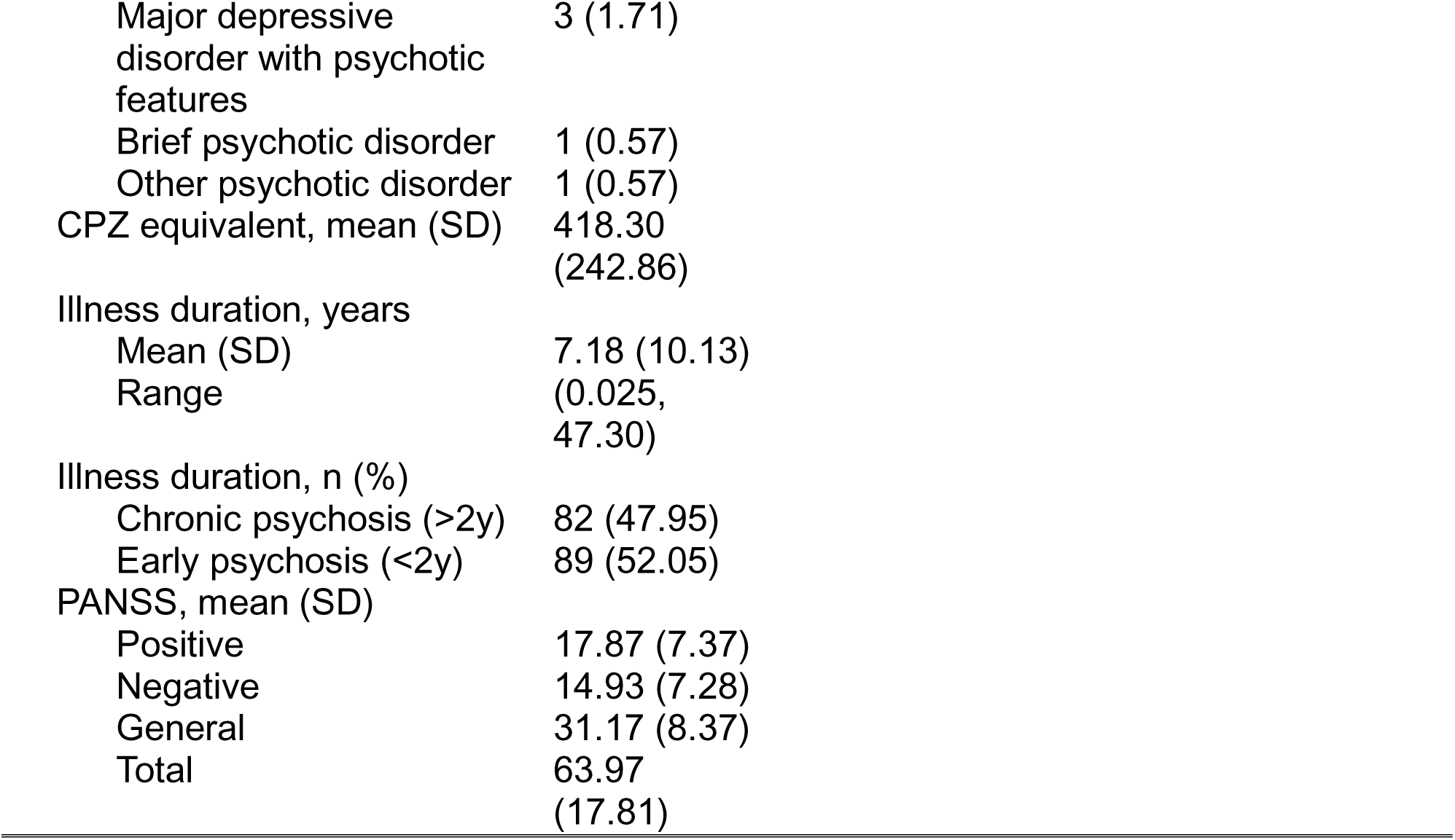
Baseline characteristics of study sample. CPZ, chlorpromazine; SD, standard deviation. *: 0.01<p<0.05, **: 0.001<p<0.01, ***: p<0.001.

| | Psychosis<br>($n=175$) | Control<br>($n = 161$) | P-value | Test |
| --- | --- | --- | --- | --- |
| **Demographics** |  |  |  |  |
| Age in years, mean (SD) | 28.81<br>(10.68) | 29.22<br>(9.96) | 0.715 | T-test |
| Sex, female, $n$ (%) | 57 (32.57) | 61 (37.89) | 0.365 | Chi-square |
| Race, $n$ (%) | | | 0.1712 | Chi-square |
| White | 111 (62.43) | 117 (72.67) |  |  |
| Black or African American | 52 (29.71) | 34 (21.11) |  |  |
| Other | 12 (6.86) | 10 (6.21) |  |  |
| **Tobacco use history** |  |  |  |  |
| Ever-use, $n$ (%) | 106 (60.57) | 31 (19.25) | 3.249e-14*** | Chi-square |
| Current-use, $n$ (%) | 75 (42.86) | 17 (10.56) | 7.503e-11*** | Chi-square |
| **Psychotic illness history** |  |  |  |  |
| Diagnosis, $n$ (%) | | | | |
| Schizophrenia | 62 (35.43) |  |  |  |
| Schizophreniform | 54 (30.86) |  |  |  |
| Bipolar disorder with psychotic features | 31 (17.71) |  |  |  |
| Schizoaffective disorder | 23 (13.14) |  |  |  |

|  |  |
| --- | --- |
| Major depressive disorder with psychotic features | 3 (1.71) |
| Brief psychotic disorder | 1 (0.57) |
| Other psychotic disorder | 1 (0.57) |
| CPZ equivalent, mean (SD) | 418.30<br>(242.86) |
| Illness duration, years |  |
| Mean (SD) | 7.18 (10.13) |
| Range | (0.025, 47.30) |
| Illness duration, n (%) |  |
| Chronic psychosis (>2y) | 82 (47.95) |
| Early psychosis (<2y) | 89 (52.05) |
| PANSS, mean (SD) |  |
| Positive | 17.87 (7.37) |
| Negative | 14.93 (7.28) |
| General | 31.17 (8.37) |
| Total | 63.97<br>(17.81) |

### Higher DMN Connectivity in Psychosis Predicts Higher Likelihood of Lifetime Tobacco Use

We observed a significant diagnosis x DMN connectivity interaction for lifetime tobacco use (p = 0.0281, coefficient = 0.457, OR = 1.579, 95% CI = [1.063, 2.411]; Figure 1, Supplementary Table 2), controlling for age, sex, medication, and scanning protocol. Diagnosis influenced the relationship between DMN connectivity and tobacco use; in individuals with psychosis, higher DMN connectivity was associated with higher odds of tobacco use, while in controls, higher DMN connectivity was associated with lower likelihood of tobacco use. We did not observe main or diagnosis interaction effects of SN or DMN-SN connectivity on likelihood of lifetime tobacco use (both p > 0.1; Supplementary Table 2).

**Figure 1.**
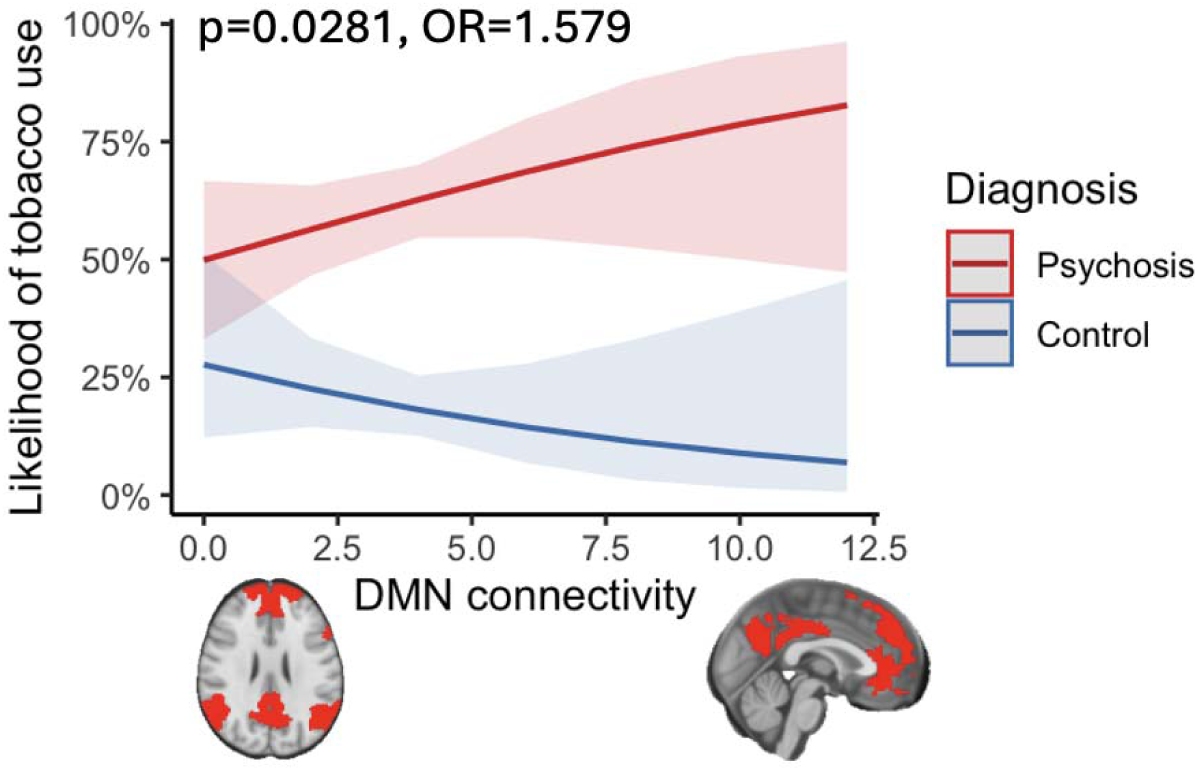
To investigate how diagnosis influence the relationship between network connectivity and likelihood of lifetime tobacco use, we performed logistic regression to model tobacco use using diagnosis-by-connectivity interaction. We observed a significant diagnosis x DMN connectivity interaction for lifetime tobacco use (p = 0.0281, coefficient = 0.457, OR = 1.579, 95% CI = [1.063, 2.411]). Higher DMN connectivity was associated with higher odds of lifetime tobacco use in psychosis, but with lower odds in controls. DMN, default mode network.

### Altered Connectivity in Some Within-DMN and DMN-SN Edges Predicts Higher Likelihood of Lifetime Tobacco Use

To identify the most impactful connectivity edges in influencing tobacco use, we trained a logistic LASSO regression model to predict likelihood of lifetime tobacco use using diagnosis x edge-level connectivity, controlling for age, sex, medication, scanner, and scanning protocol. Model performance was AUC = 0.799 (95% CI = [0.711, 0.887]), sensitivity = 0.875, specificity = 0.617 (Supplementary Figure 3) when evaluated on the held-out testing dataset. Bootstrapping the model yielded four parameters with stable nonzero regression coefficients (beta), suggesting statistical significance: age (beta = 0.006, 95% CI = [0.004, 0.012]), diagnosis (beta = −0.143, 95% CI = [−0.286, −0.003]), LLP_DMN_–LaPFC_SN_ connectivity (beta = −0.007, 95% CI = [−0.015, - 0.001]), and diagnosis x LLP_DMN_–RLP_DMN_ connectivity (beta = 0.012, 95% CI = [0.0001, 0.025]. Lower LLP_DMN_–LaPFC_SN_ connectivity was related to higher odds of tobacco use irrespective of diagnosis. Higher LLP_DMN_–RLP_DMN_ connectivity increases likelihood of tobacco use only in psychosis but not in control (Supplementary Table 3).

To validate the results of LASSO regression, we performed post-hoc analysis to compare LLP_DMN_–RLP_DMN_ and LLP_DMN_–LaPFC_SN_ connectivity by tobacco use history. Post-hoc t-test showed that, only in the psychosis group, lifetime tobacco users had higher LLP_DMN_–RLP_DMN_ connectivity compared to never-users (t(142) = 2.20, p = 0.029, Figure 2A), while LLP_DMN_–RLP_DMN_ connectivity did not separate by tobacco use in the control group (p > 0.1). Across the entire sample, LLP_DMN_–LaPFC_SN_ connectivity was more negative (i.e., more anticorrelated) in lifetime tobacco users compared to never-users (t(290) = −2.69, p = 0.00755, Figure 2B), while the connectivity did not differ by diagnosis (p > 0.1).

**Figure 2.**
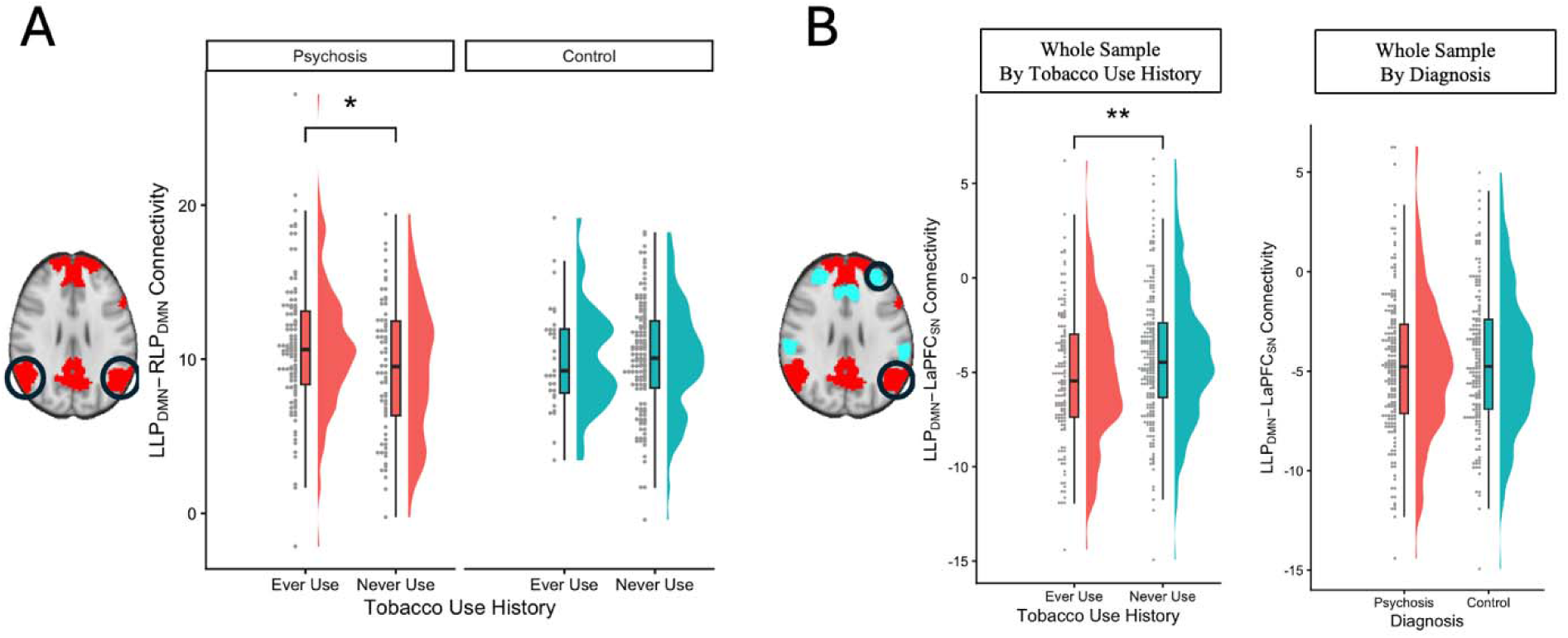
We performed post-hoc analysis to validate the most impactful connectivity edges influencing tobacco use, as selected by the LASSO regression model. (A) Only in the psychosis group did lifetime tobacco users have higher LLP_DMN_–RLP_DMN_ connectivity compared to never-users (p = 0.029), but no difference between lifetime-and never-use was observed in control participants (p > 0.1). (B) Across the entire sample, LLP_DMN_–LaPFC_SN_ connectivity was more negative (i.e., more anticorrelated) in lifetime tobacco users compared to never-users (p = 0.0097), while the connectivity did not differ by diagnosis (p > 0.1). DMN, default mode network; LaPFC, left anterior prefrontal cortex; LLP, left lateral parietal; RLP, right lateral parietal; SN, salience network.

### Daily Cigarette Use is Correlated with DMN Connectivity to the Motor Cortex and Insula

Among individuals with lifetime tobacco use, brainwide connectivity with LLP_DMN_ did not differ between psychosis and controls. However, among all participants with lifetime tobacco use, brainwide connectivity analysis identified two clusters in the bilateral motor cortices (peak voxels located at MNI [+54, −4, +28] and [−64, −6, +28]), where higher LLP_DMN_–cluster connectivity was associated with higher daily cigarette use (Figure 3). There was no significant diagnosis x smoking interaction effect on connectivity (Supplementary Table 4).

**Figure 3.**
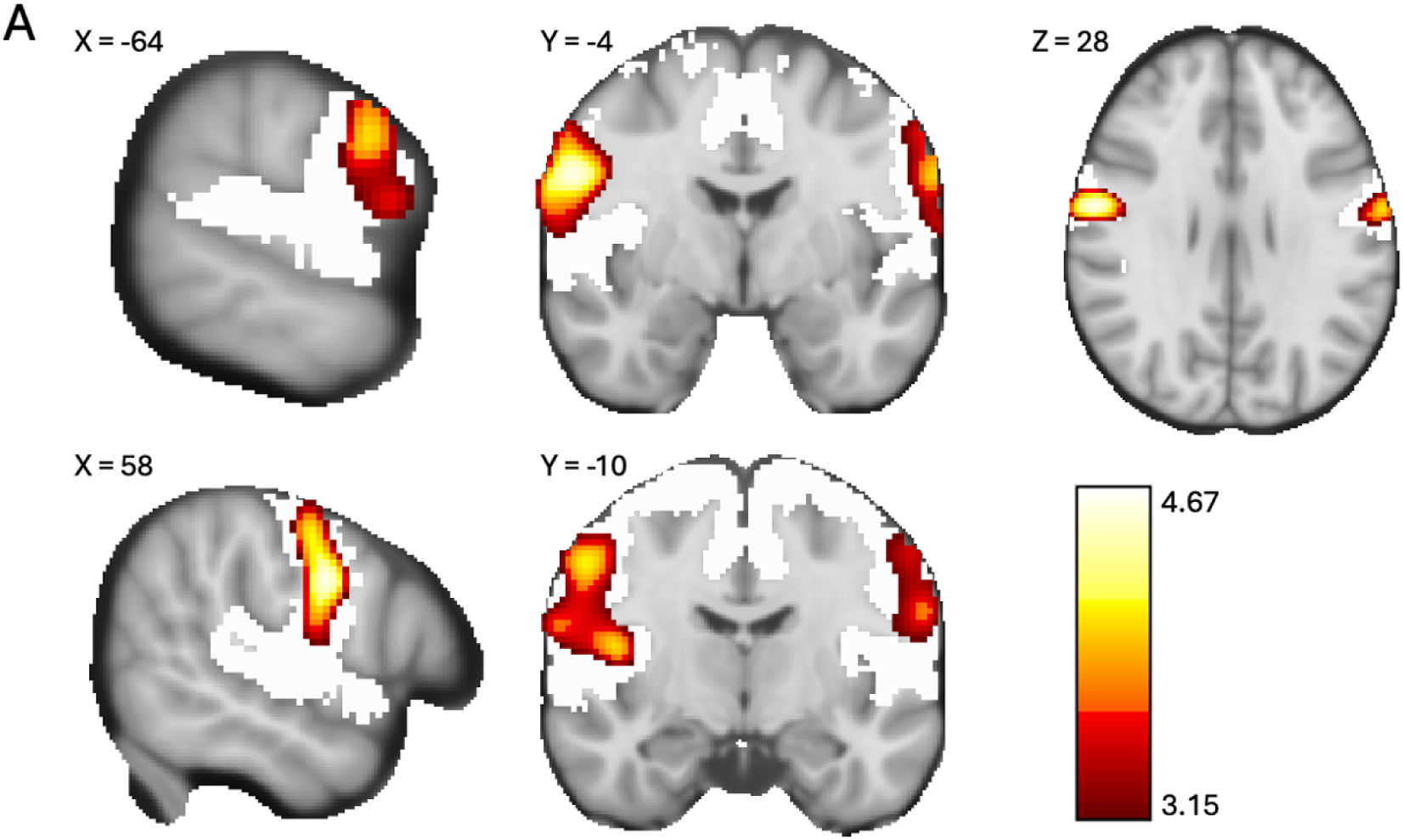
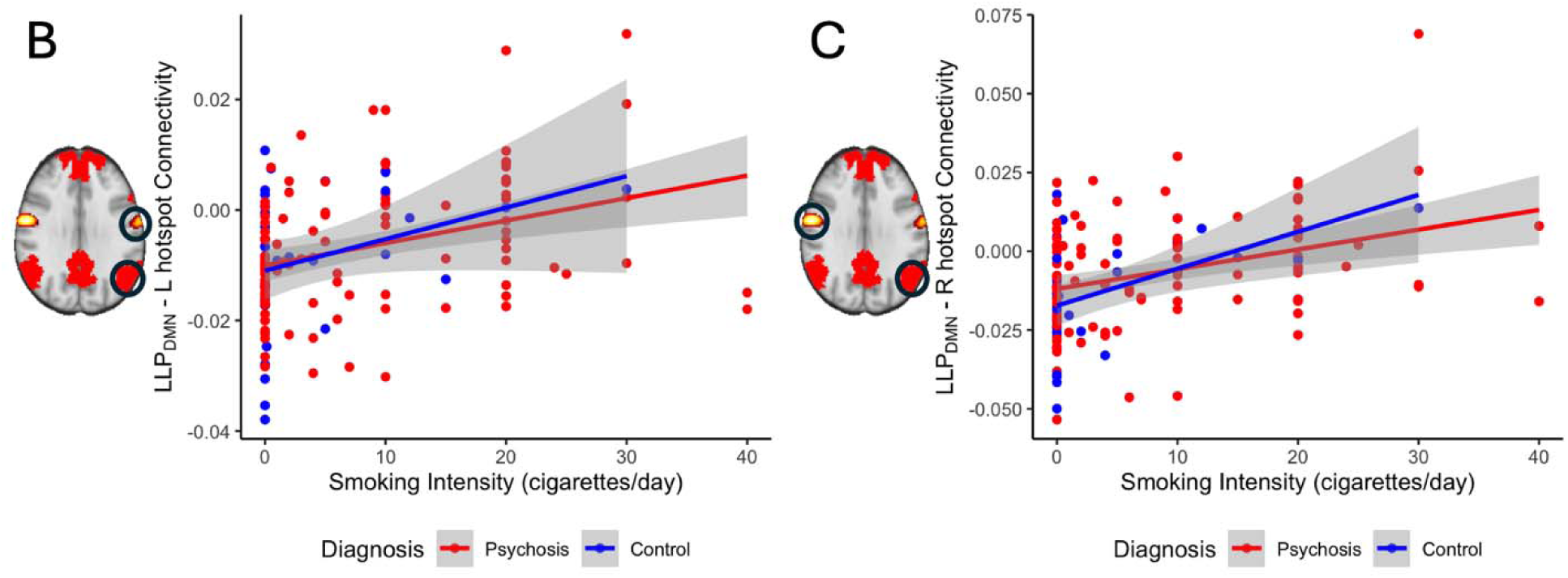
We performed brainwide analysis to identify clusters whose connectivity to LLP_DMN_ correlated with daily cigarette use. Among all participants with lifetime tobacco use, we found two clusters in the bilateral motor cortices, where higher connectivity to LLP_DMN_ correlated with higher daily cigarette use. (A) Brainwide discovery clusters (red) fell within the sensorimotor system (white). (B-C) Post-hoc visualization confirmed the relationship between current daily cigarette use and LLP_DMN_–hotspot connectivity among all ever-users of tobacco for the hotspot on the left (B) and on the right (C). Color bar displays T-statistic. Displayed map uses cluster-level threshold of p<0.05. DMN, default mode network; LLP, left lateral parietal.

Clusters spanned both precentral and postcentral gyri and encompassed portions of lower and upper face M1 ROIs (Supplementary Figure 4A)^25^ without overlapping with lower or upper face primary sensory cortex (S1) ROIs (Supplementary Figure 4B). Clusters overlapped with right posterior insula (Supplementary Figure 5A). Using a right posterior insula seed,^27^ post-hoc analysis showed a significant positive correlation between higher daily cigarette use and higher LLP_DMN_ – right posterior insula connectivity (r = 0.2032, t(135) = 2.411, p = 0.01725; Supplementary Figure 5B) among all lifetime tobacco users, with no diagnosis-by-tobacco interaction (p > 0.1). When analyzing this relationship within participants with current tobacco use, we observed the same positive association between higher daily cigarette use and higher LLP_DMN_ – right posterior insula connectivity (r = 0.2232, t(90) = 2.1727, p = 0.03243), with trend-level diagnosis x tobacco interaction in current use (estimate = −7.9e-04, SE = 4.2e-04, t = - 1.769, p = 0.0806; Supplementary Figure 5C). In participants with current tobacco use, higher daily cigarette use correlated with stronger LLP_DMN_ – right posterior insula connectivity only in control participants (r = 0.6639, t(15) = 3.4386, p = 0.00366) but not in psychosis (Supplementary Table 5).

## Discussion

Using a large sample of individuals with psychosis spectrum disorders, we conducted both ROI-based and brainwide analyses to investigate how psychosis diagnosis influenced the relationship between connectivity and tobacco use. In the ROI-based analysis, we found a psychosis-specific relationship between DMN connectivity and lifetime tobacco use. In the brainwide exploratory analysis, we identified dose-dependent relationships between current cigarette use and DMN-insula and DMN-somatomotor connectivity among people with lifetime tobacco use, irrespective of diagnosis.

### Psychosis-Specific Relationship between Altered DMN Connectivity and Tobacco Use

We found that higher DMN connectivity predicts a higher likelihood of lifetime tobacco use only in individuals with psychotic disorders but not in the control sample. When we investigated individual within-DMN and DMN-SN connections, we observed that higher connectivity between the left and right parietal DMN nodes predicted lifetime tobacco use only in individuals with psychosis, whereas stronger anti-correlation between the LLP_DMN_ and LaPFC_SN_ nodes was linked to lifetime tobacco use irrespective of diagnosis.

Our results provide confirmatory evidence supporting the role of abnormal DMN connectivity in driving tobacco use in psychosis. In a data-driven analysis with individuals with schizophrenia, Ward et al identified that hyperconnectivity of DMN regions had the strongest link with cigarette use across the connectome, and DMN hyperconnectivity was normalized by acute nicotine administration only in schizophrenia.^9^ In another study, Ward et al further confirmed this relationship between DMN connectivity and tobacco use in a larger sample of individuals with psychotic spectrum disorders, reporting lower DMN connectivity in current smokers with psychosis compared to former or never smokers.^21^ However, while previous studies have reported higher DMN connectivity in individuals with psychosis and in tobacco users separately,^8–11,15,21^ they have been underpowered to find a significant interactive effect. Our findings begin to fill this gap by suggesting that DMN modulation may be a more effective target for tobacco use interventions among people with psychosis. Recent work using DMN-targeted TMS on smokers without psychosis did not affect craving.^22^ However, previous work has shown that DMN-targeted TMS does modulate DMN connectivity and reduce craving in people with psychosis,^21^ suggesting diagnostic specificity in treatment outcomes. Our findings linking bilateral parietal DMN connectivity to tobacco use highlight this connectivity edge as a target for psychosis-specific interventions for tobacco use.

### Altered DMN-Insula and DMN-Somatomotor Connectivity in Cigarette Use

In our brainwide exploratory analysis, we also observed novel dose-dependent relationships between current cigarette use and LLP_DMN_ connectivity to right posterior insula and bilateral motor cortex among people with lifetime tobacco use, irrespective of diagnosis. The insula finding is not surprising, as the insula is implicated in sensory, affective, and cognitive processing^29^ and plays a role in reward anticipation.^30^ Our findings are consistent with the insula’s established role in nicotine addiction among individuals without psychosis.^31^ In observational studies, people who smoked cigarettes experienced disruption in smoking addiction following insula stroke,^32,33^ and cue-induced insula reactivity prior to smoking cessation predicted ability to maintain tobacco abstinence.^34^ Animal models have also shown that inactivating the posterior insula decreased tobacco-seeking behavior in rats.^35,36^ Given the broad evidence for the role of the insula in nicotine dependence, it has been a target for neuromodulatory interventions. Deep TMS targeting the bilateral DLPFC and insula is FDA-cleared for smoking cessation.^19^ Furthermore, in people without psychosis, higher insula-DMN connectivity has been associated with cue-induced tobacco craving,^37,38^ and this connectivity was modulated by both varenicline and tobacco patch.^39^ Our findings were consistent with the established link between insula-DMN connectivity in tobacco use among individuals without psychosis. Interestingly, this relationship was only present in control participants in our analysis. However, a larger sample size is needed before inferring any differential effect that psychosis may have on the relationship between insula-DMN connectivity and tobacco use.

Compared to the insula, the role of the motor system in substance use has yet been as extensively explored. Casartelli and Chiamulera proposed the “motor cognition” hypothesis, where the motor system is involved in the psycho-pathophysiology of substance use disorders by establishing automatized/compulsive behaviors in brain regions encoding skill learning and action meaning.^40^ Existing neuroimaging studies have investigated cue-induced sensorimotor system response, showing stronger cue-induced activity in superior sensorimotor cortex,^41^ and more severe tobacco dependence correlated with higher activity in regions encoding action knowledge^42^, tool use skills,^42^ motor preparation, ^43^ and visuospatial attention.^43^ However, no study has reported resting-state somatomotor system connectivity relationships with tobacco use. Our finding of resting-state DMN-motor cortex connectivity is surprising, since our participants were not responding to smoking cues or instructed to think about smoking. It is also worth noting that our discovery cluster overlapped with lower face M1, facial regions heavily involved in smoking behavior. Unlike our insula-DMN finding, we did not observe any diagnosis x smoking interaction on the relationship between DMN-somatomotor connectivity and smoking intensity. Though we could not infer specific conclusions from our findings, motor cognition – as facilitated by DMN-somatomotor connectivity – may be a promising area for future research on substance use, and findings could potentially apply to both individuals with and without psychosis.

### Strengths and Limitations

Our analysis has several strengths. First, we used a large sample of individuals with psychotic disorders and non-clinical control participants who reported their tobacco use history. We used analytic approaches to test hypotheses that had previously been observed only in small samples. Second, our sample included participants with a wide range of psychotic illness diagnoses, durations of illness, and subtypes (i.e., both affective and nonaffective). However, our analysis has several limitations. First, the cross-sectional nature of these relationships cannot confirm causality. To this end, we have observed in previous interventional experiments that 1) acute tobacco administration reduces DMN hyperconnectivity only in psychosis participants;^9^ and 2) DMN-targeted TMS reduces DMN connectivity and reduces craving.^21^ These studies suggest that using interventions to reduce DMN connectivity are potentially effective to reduce tobacco use among people with psychotic disorders. Another limitation of our analysis that our psychosis and control groups were not matched in tobacco use due to the low prevalence of tobacco use in people without psychosis at large. These rates of tobacco use roughly match population-level rates in individuals with and without psychotic disorder diagnoses,^3^ but provide a potential confounder to our results. Additionally, there is a lack of granularity in our tobacco use data. Future studies should obtain information both combustible and noncombustible tobacco use, craving and withdrawal, increase recruitment of nonsmokers without psychosis, and test interventions to reduce DMN connectivity and measure effects on tobacco use, craving, and withdrawal.

In conclusion, this is the first analysis to provide evidence that LLP_DMN_ connectivity relates to lifetime tobacco use in psychotic disorders, supporting the LLP_DMN_ as a psychosis-specific brain target for tobacco cessation interventions. Given the high prevalence of tobacco use by people with psychotic disorders and the inferiority of current tobacco cessation interventions, this finding suggests a promising target for future highly effective interventions to save the decades of life lost to tobacco-related illness in this vulnerable population.

## Data Availability

All data produced in the present study are available upon reasonable request to the authors.

## Author Contributions

HBW conceived and designed the work. Acquisition and analysis of data was completed by YB, AK, NW, SH, and SV. Interpretation, drafting, and revision for publication were completed by YB and HBW. All authors gave final approval for publication and agree to be accountable for all aspects of the work.

## Acknowledgments

This work was supported by National Institutes of Health (NIH) grants R01MH102266 to Dr. Woodward, R01MH070560 to Dr. Heckers, K23MH126313 to Dr. Sheffield, and K23DA059690 to Dr. Ward; by Vanderbilt Institute for Clinical and Translational Research grant VR71021 to Mr. Kittleson; and by Vanderbilt University School of Medicine Medical Scholars Program Fellowship to Ms. Bai.

