## Supplement for "Parietal Default Mode Network Connectivity is Associated with Tobacco Use in Psychosis"

**Supplemental Methods**

*Participants*

Data came from a repository of 336 individuals with psychotic disorders and non-clinical controls with complete neuroimaging and behavioral data who participated in one of four neuroimaging projects conducted at Vanderbilt University Medical Center (VUMC, CT00762866; 1R01MH070560; 1R01MH102266; VR71021). All studies were approved by the Vanderbilt Institutional Review Board, and all individuals provided written informed consent prior to participating in the studies. Exclusion criteria across the three studies were: age under 16/18 years or over 65 years (55 years in 1R01MH102266, 50 years in VR71021); estimated premorbid IQ less than 70 based on the Wechsler Test of Adult Reading; history of significant head trauma, medical illness, or central nervous system disorder; pregnancy or lactation; substance abuse within the last 1 month for psychosis participants (3 months in 1R01MH102266); and MRI contraindicators. VR71021 had additional exclusion criteria of inability to communicate in English and lacking normal or corrected-to-normal vision.

*MRI Acquisition*

For a subset of individuals (n=28), a cognitive task preceded the resting-state scan. In this task, participants are instructed to count their heart beats without felling for their pulse for a certain amount of time.

Scan parameters are shown in table below.

| **Scan Types** | **TR (msec)** | **TE (msec)** | **Flip Angle** | **Voxel Size (mm^3^)** | **Volumes** | **FOV mm (x)** | **Slice Thickness (mm)** |
| --- | --- | --- | --- | --- | --- | --- | --- |
| T1 | 8 | 4 | 5 | 1 | 1 | 170 | 1 |
|  | 9 | 5 | 8 | 1 | 1 | 170 | 1 |
| Resting-State fMRI | 2000 | 35 | 79 | 3 | 203 | 240 | 4 |
|  | 2000 | 28 | 90 | 3 | 210 | 240 | 3.2 |
|  | 2000 | 28 | 90 | 3 | 203 | 240 | 3.2 |
|  | 2000 | 25 | 90 | 3 | 300 | 240 | 3 |

*Calculation of Network- and Edge-Level Connectivity Values*

We calculated network-level connectivity for within-DMN, within-SN, and between-DMN-SN connectivity values, and edge-level connectivity for all connections (edges) between LLP_DMN_ and other DMN and SN nodes. Whole-network connectivity values were calculated using nine standard nodes of default mode network (DMN) and seven standard nodes of salience network (SN) using MNI coordinates listed in the follow table:

| **Network** | **Node** | **MNI Coordinate** |
| --- | --- | --- |
| DMN | Posterior cingulate/precuneus | [0 -52 27] |
|  | Medial prefrontal | [-1 54 27] |
|  | Left lateral parietal | [-46 -66 30] |
|  | Right lateral parietal | [49 -63 33] |
|  | Left inferior temporal | [-61 -24 -9] |
|  | Right inferior temporal | [58 -24 -9] |
|  | Medial dorsal thalamus | [0 -12 9] |
|  | Right posterior cerebellum | [-25 -81 -33] |
|  | Left posterior cerebellum | [25 -81 -33] |
| SN | Dorsal anterior cingulate | [0 21 36] |
|  | Left anterior prefrontal | [-35 45 30] |
|  | Right anterior prefrontal | [32 45 30] |
|  | Left insula | [-41 3 6] |
|  | Right insula | [41 3 6] |
|  | Left lateral parietal | [-62 -30 12] |
|  | Right lateral parietal | [59 -27 15] |

*Supplemental statistical analyses*

We investigated group differences in network connectivity by diagnosis (psychosis VS control), tobacco use status (lifetime-use VS never-use), and psychotic illness duration (chronic psychosis [> 2 years], early psychosis [< 2 years], or control). Welch’s two-sample t-tests were used to compare difference in connectivity between dichotomous variables, including diagnosis and tobacco use status. One-sample ANOVAs with Bonferroni-adjusted pairwise comparisons were used to compare difference in connectivity based on psychotic illness duration. Robust variance estimator was used for ANOVAs to account for heteroskedasticity.

We investigated group differences in psychosis symptom severity by tobacco use status (lifetime-use VS never-use) using Welch’s two-sample t-tests. Psychosis symptoms were measured using the Positive and Negative Syndrome Scale (PANSS). The Marder factor analysis was used to calculate positive, negative, and general psychopathology subscores.

**Supplemental Results**

No significant group differences in network connectivity (within-DMN, within-SN, or DMN-SN) were observed between psychosis and controls, or between people with lifetime or no use of tobacco (all p > 0.1; Supplementary Figure 1, Supplementary Table 1).

Comparing network connectivity by illness duration, we observed 1) lower DMN connectivity in early psychosis compared to chronic psychosis (estimate = 0.696, SE = 0.254, t(327) = 2.744, FWERp = 0.0192) and to controls (estimate = 0.550, SE = 0.179, t(327) = 3.067, FWERp = 0.0070), 2) no difference in SN connectivity by illness duration, and 3) more negative DMN-SN connectivity (i.e., more anticorrelated) in chronic psychosis compared to early psychosis (estimate = -0.4686, SE = 0.179, t(327) = -2.619, FWERp = 0.0277; Supplementary Figure 2, Supplementary Table 1).

Lifetime tobacco use was not associated with psychotic symptoms. Psychosis participants with lifetime tobacco use did not differ from those with never-use in terms of PANSS total, positive, negative, or general psychopathology scores (all p > 0.1).

**Supplemental Figures**

Supplementary Figure 1

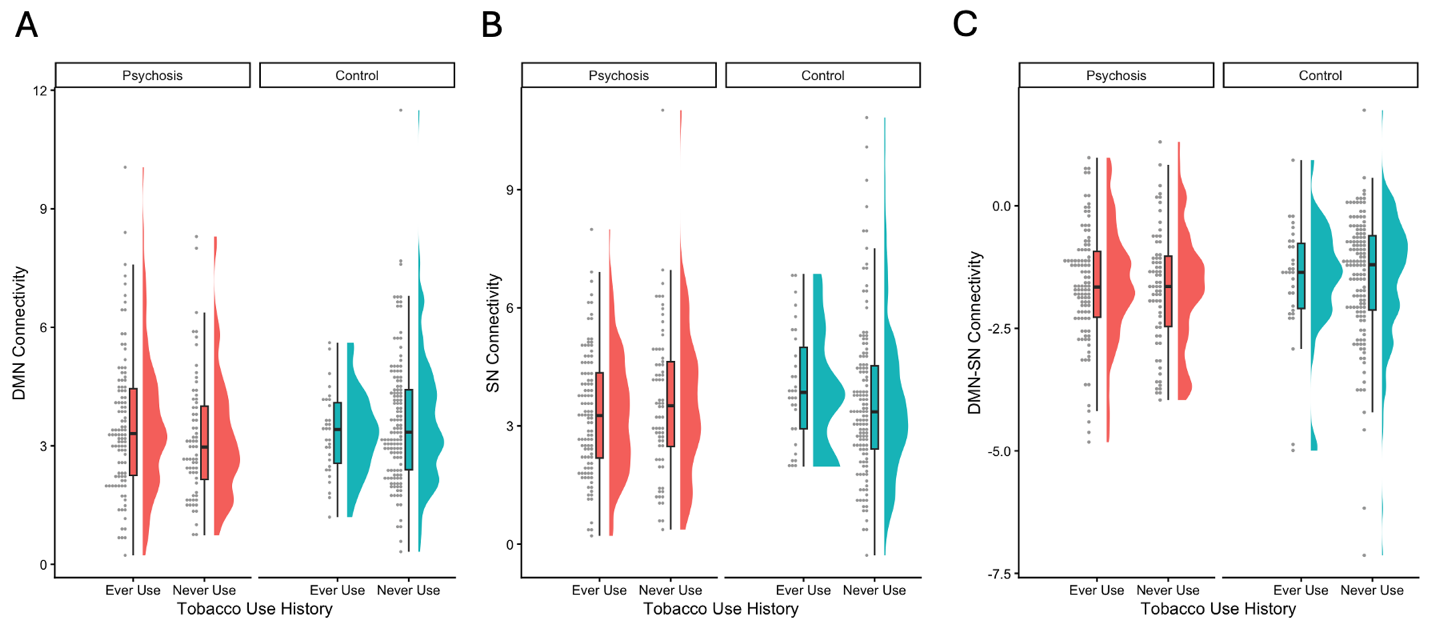

Supplementary Figure 1. Network-level connectivity analysis results. Within DMN (A), Within SN (B), DMN-SN (C) network connectivity did not differ by psychosis diagnosis or by tobacco use history (all p > 0.1).

Supplementary Figure 2
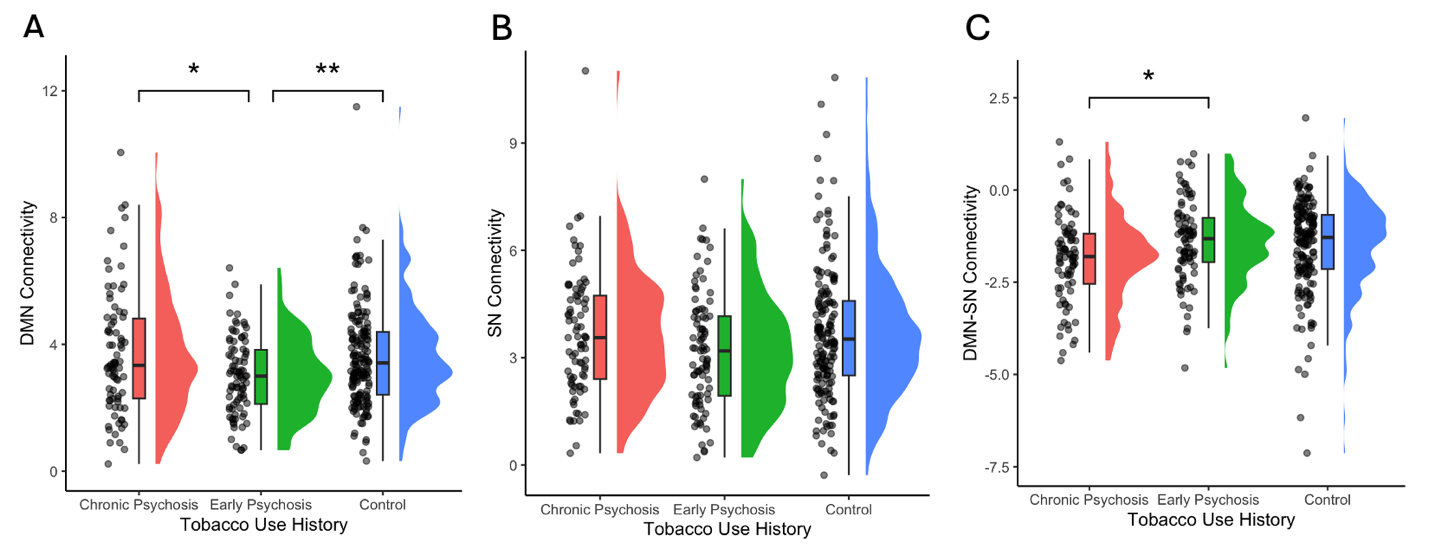

Supplementary Figure 2. (A) DMN connectivity was lower in early psychosis compared to control participants (FWERp = 0.0192) and to chronic psychosis (FWERp = 0.0070). (B) SN connectivity did not differ by psychotic illness duration. (C) DMN-SN connectivity was more anti-correlated (stronger inhibition) in chronic psychosis compared to early psychosis (FWERp = 0.0277) but not to control (p > 0.05). DMN, default mode network; SN, salience network. *: 0.01<p<0.05, **: 0.001<p<0.01, ***: p<0.001.

Supplementary Figure 3

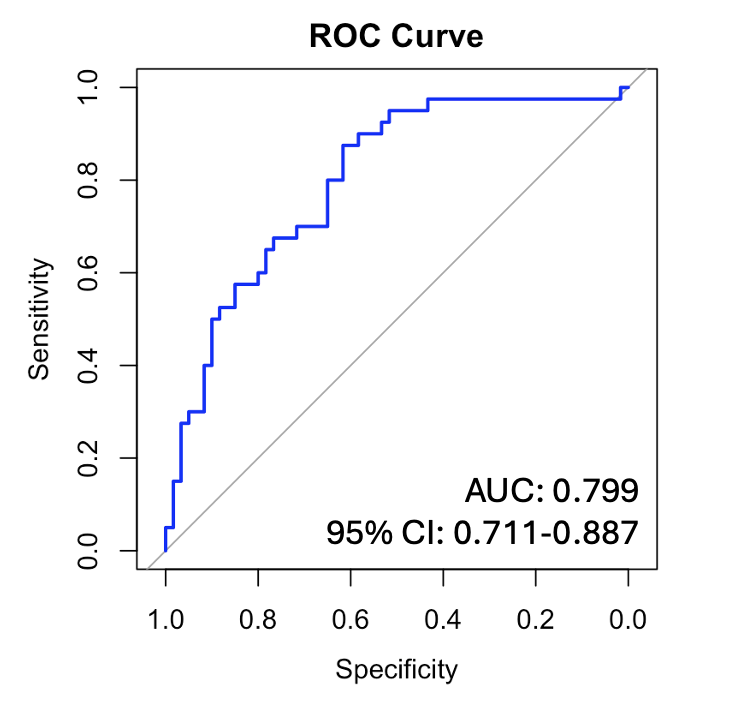

Supplementary Figure 3. Response-operator curve of refitted model using important predictors obtained from LASSO regression with bootstrapping. Refitted model achieved AUC of 0.799 (95% CI: [0.711, 0.887]). AUC, area under curve; CI, confidence interval; ROC, response-operator curve.

Supplementary Figure 4

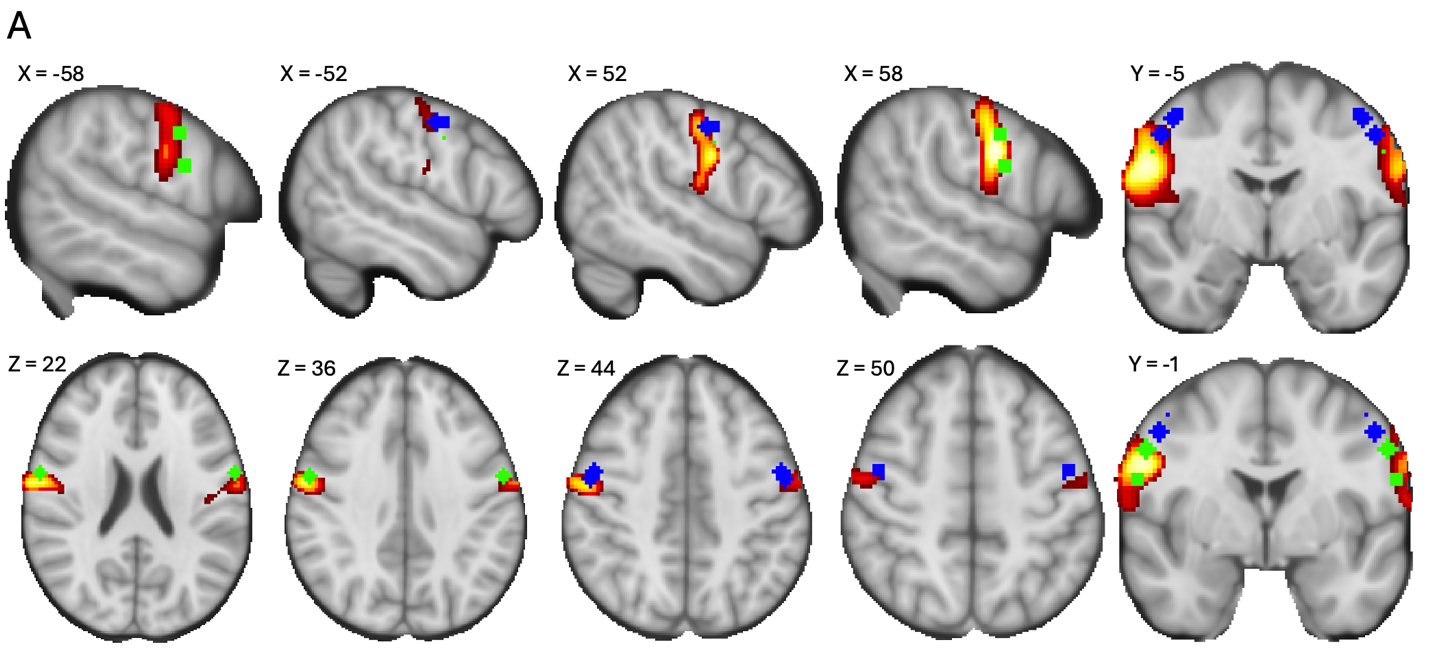

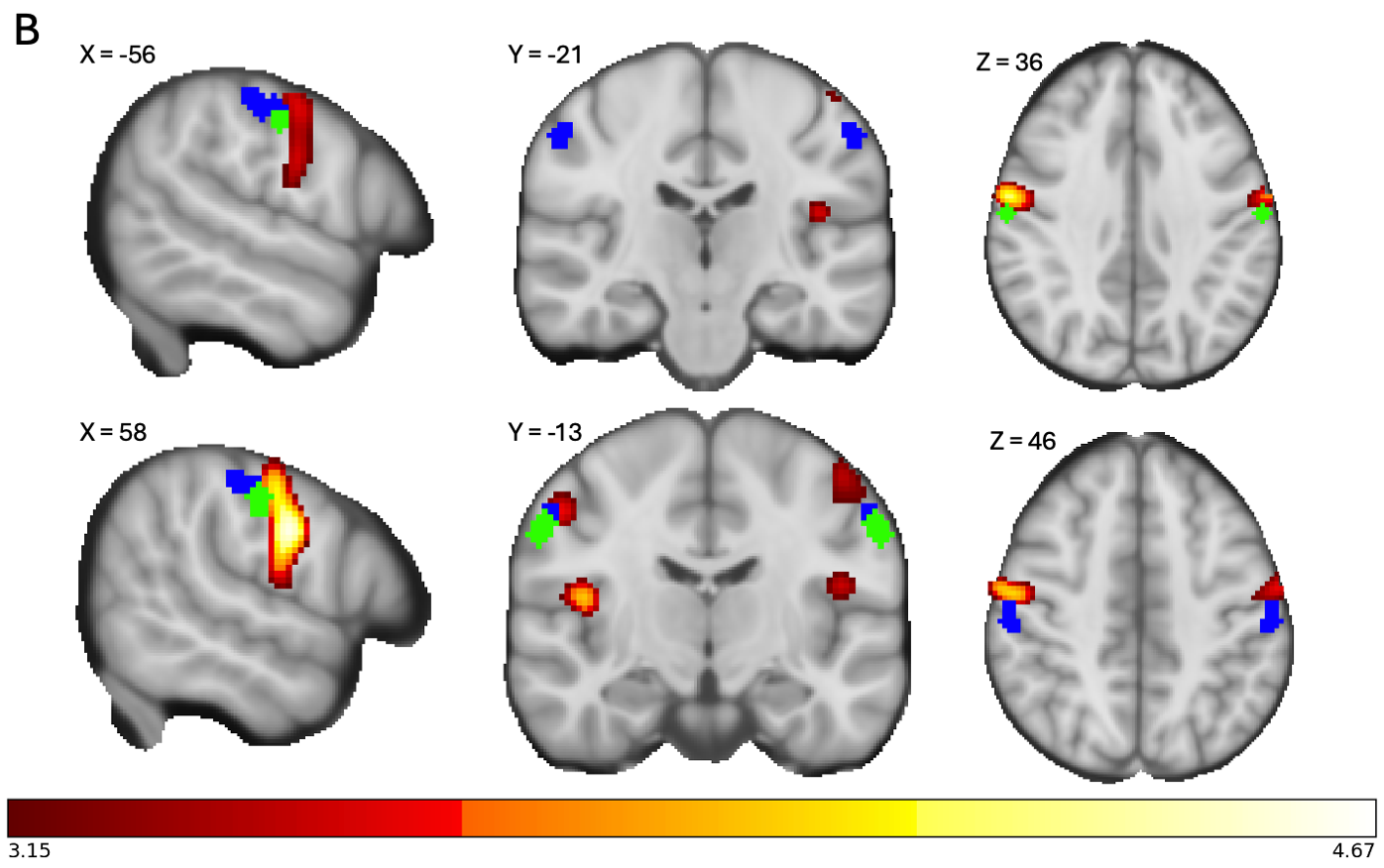

Supplementary Figure 4. (A) Brainwide discovery clusters (red) partially overlapped with lower face M1 (green) and upper face M1 (blue) ROIs, but (B) did not overlap with lower face S1 (green) or upper face S1 (blue) ROIs. Color bar displays T-statistic. Displayed map is thresholded at p<0.01. X, Y, and Z, indicated MNI values at which the images were obtained. M1, primary motor cortex; S1, primary sensory cortex; ROI, region of interest.

Supplementary Figure 5

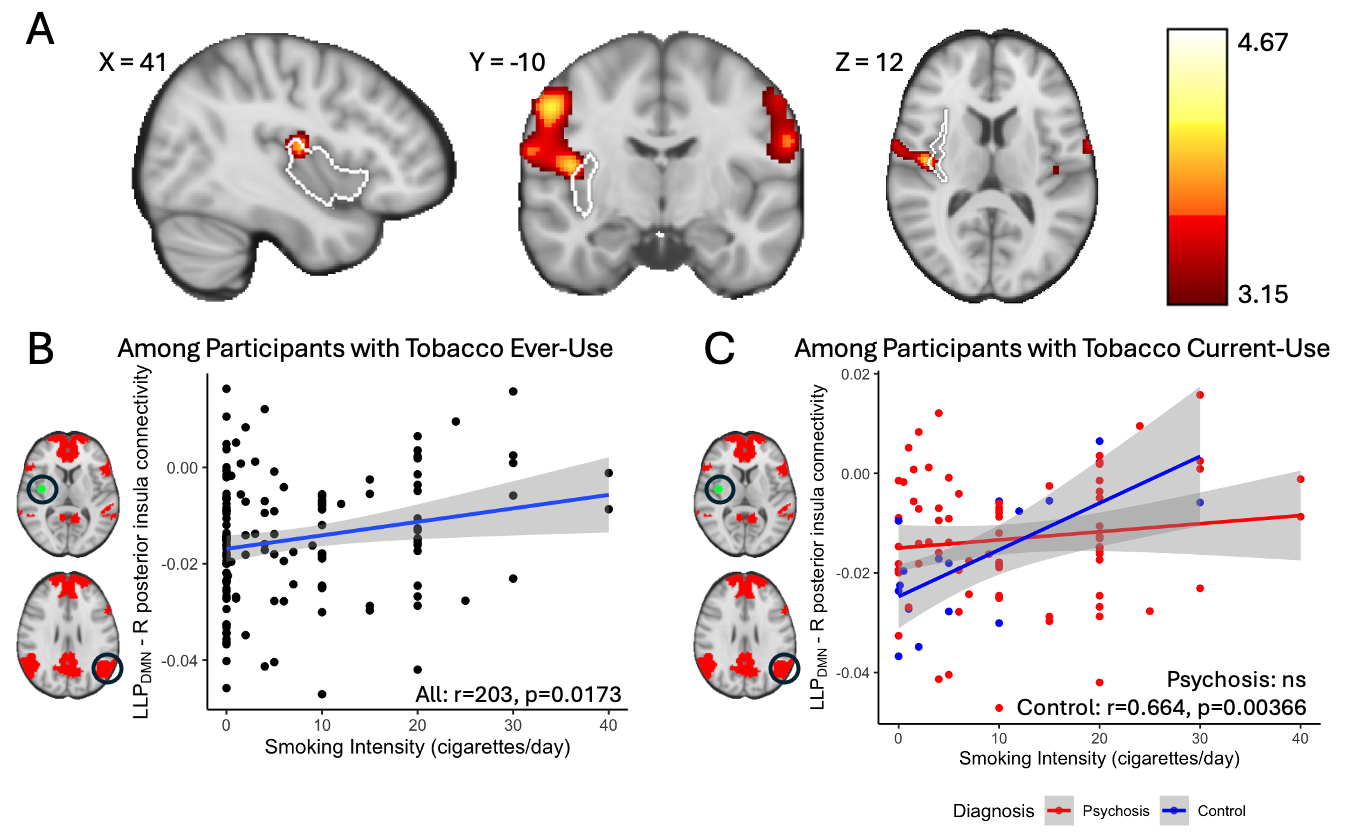

Supplementary Figure 5. A) Brainwide discovery clusters (red) overlapped with right posterior insula mask (white). (B) Higher smoking intensity correlated with higher LLP_DMN_ connectivity to right posterior insula (green) in participants with ever-use. (C) In participants with current-use, higher smoking intensity correlated with higher LLP_DMN_ connectivity to right posterior insula only in controls but not in psychosis, with trend-level interaction (p = 0.0806). Color bar displays T-statistic. Displayed map is thresholded at p<0.01. DMN, default mode network; LLP, left lateral parietal; r, Pearson’s coefficient.

**Supplemental Tables**

**Supplementary Table 1**

| Network connectivity difference by diagnosis (Welch’s two sample t-tests) | | | | | |
| --- | --- | --- | --- | --- | --- |
| Network | Psychosis (n=175)  Mean (SD) | Control (n=161)  Mean (SD) | DoF | t-value | p-value |
| DMN | 3.40 (1.69) | 3.56 (1.54) | 332 | 0.868 | 0.386 |
| SN | 3.43 (1.69) | 3.68 (1.89) | 322 | 1.29 | 0.199 |
| DMN-SN | -1.64 (1.18) | -1.47 (1.27) | 325 | 1.28 | 0.202 |

| Network connectivity difference by tobacco use history (Welch’s two-sample t-tests) | | | | | |
| --- | --- | --- | --- | --- | --- |
| Network | Ever-use (n=137)  Mean (SD) | Never-use (n=199)  Mean (SD) | DoF | t-value | p-value |
| DMN | 3.49 (1.62) | 3.47 (1.62) | 291 | 0.137 | 0.891 |
| SN | 3.48 (1.55) | 3.60 (1.94) | 326 | -0.671 | 0.503 |
| DMN-SN | -1.60 (1.17) | -1.54 (1.27) | 304 | -0.422 | 0.673 |

| Network connectivity difference by illness duration (ANOVA) | | | | | | |
| --- | --- | --- | --- | --- | --- | --- |
|  | Chronic psychosis  (n=82)  Mean (SD) | Early psychosis (n=89)  Mean (SD) | Control (n=161)  Mean (SD) | DoF | F | p-value |
| DMN | 3.70 (1.95) | 3.01 (1.22) | 3.56 (1.54) | (2, 327) | 6.156 | 0.00238** |
| SN | 3.64 (1.74) | 3.19 (1.62) | 3.68 (1.89) | (2, 329) | 2.629 | 0.0740 |
| DMN-SN | -1.86 (1.20) | -1.39 (1.11) | -1.47 (1.27) | (2, 327) | 3.856 | 0.0221* |

| Post-hoc multiple comparisons by illness duration | | | | | | |
| --- | --- | --- | --- | --- | --- | --- |
|  | | Estimate | SE | DoF | t-value | FWERp |
| DMN | |  |  |  |  |  |
|  | Chronic psychosis – Control | 0.146 | 0.249 | 327 | 0.586 | 1.0000 |
|  | Chronic psychosis – Early psychosis | 0.696 | 0.254 | 327 | 2.744 | 0.0192* |
|  | Early psychosis – Control | 0.550 | 0.179 | 327 | 3.067 | 0.0070** |
| SN | |  |  |  |  |  |
|  | Chronic psychosis – Control | -0.0389 | 0.245 | 329 | -0.159 | 1.0000 |
|  | Chronic psychosis – Early psychosis | 0.4552 | 0.260 | 329 | 1.752 | 0.2421 |
|  | Early psychosis – Control | 0.4942 | 0.229 | 329 | 2.161 | 0.0942 |
| DMN-SN | |  |  |  |  |  |
|  | Chronic psychosis – Control | -0.3847 | 0.167 | 327 | -2.303 | 0.0657 |
|  | Chronic psychosis – Early psychosis | -0.4686 | 0.179 | 327 | -2.619 | 0.0277* |
|  | Early psychosis – Control | -0.0839 | 0.156 | 327 | -0.538 | 1.0000 |

Supplementary Table 1. Comparison of within- and between-network connectivity by diagnosis and by nicotine use history. DoF, degree of freedom; SD, standard deviation; SE, standard error. P-values were associated with tests for group differences in connectivity; *: 0.01<p<0.05, **: 0.001<p<0.01, ***: p<0.001.

**Supplementary Table 2**

|  | Estimate | SE | z-value | p-value | Odds ratio | 95% CI |
| --- | --- | --- | --- | --- | --- | --- |
| Intercept | -3.291 | 0.901 | -3.651 | 0.0002*** | 0.037 | [0.0061, 0.212] |
| Diagnosis | 1.567 | 0.807 | 1.942 | 0.052 | 4.792 | [0.984, 23.507] |
| DMN | -0.182 | 0.167 | -1.093 | 0.274 | 0.833 | [0.590, 1.141] |
| SN | 0.203 | 0.140 | 1.456 | 0.145 | 1.225 | [0.931, 1.615] |
| DMN-SN | -0.015 | 0.218 | -0.066 | 0.950 | 0.986 | [0.635, 1.506] |
| Diagnosis x DMN | 0.457 | 0.208 | 2.196 | 0.0281* | 1.579 | [1.063, 2.411] |
| Diagnosis x SN | -0.273 | 0.177 | -1.536 | 0.124 | 0.761 | [0.536, 1.080] |
| Diagnosis x DMN-SN | 0.206 | 0.288 | 0.715 | 0.475 | 1.229 | [0.702, 2.185] |

Supplementary Table 2. Results from logistic regression investigating the relationship between diagnosis-by-network connectivity interaction and likelihood of lifetime nicotine use. CI, confidence interval; DMN, default mode network; SE, standard error; SN, salience network. *: 0.01<p<0.05, **: 0.001<p<0.01, ***: p<0.001.

Supplementary Table 3

|  | Average beta | 95% CI | Selection frequency |
| --- | --- | --- | --- |
| **Diagnosis** | **-0.143** | **[-0.286, -0.003]** | **76.0%** |
| LLP_DMN_–posterior cingulate _DMN_ | 0 | [-0.007, 0] | 34.1% |
| LLP_DMN_–medial temporal _DMN_ | 0 | [-0.003, 0.005] | 37.7% |
| LLP_DMN_–RLP_DMN_ | 0 | [-0.007, 0] | 38.6% |
| LLP_DMN_–L inferior temporal _DMN_ | 0 | [-0.001, 0.006] | 46.4% |
| LLP_DMN_–R inferior temporal _DMN_ | 0 | [-0.005, 0.005] | 54.0% |
| LLP_DMN_–mid thalamus _DMN_ | 0 | [-0.008, 0.004] | 43.8% |
| LLP_DMN_–L posterior cerebellar _DMN_ | 0 | [-0.004, 0.007] | 58.1% |
| LLP_DMN_–R posterior cerebellar _DMN_ | 0 | [-0.003, 0.004] | 39.7% |
| LLP_DMN_–dorsal anterior cingulate _SN_ | 0.001 | [-0.006, 0.007] | 65.0% |
| **LLP_DMN_–LaPFC_SN_** | **-0.007** | **[-0.015, -0.001]** | **83.1%** |
| LLP_DMN_–RaPFC_SN_ | 0 | [-0.005, 0.006] | 36.1% |
| LLP_DMN_–LLP_SN_ | 0 | [-0.006, 0.003] | 62.6% |
| LLP_DMN_–RLP_SN_ | 0 | [-0.006, 0.004] | 45.5% |
| **Age** | **0.006** | **[0.004, 0.012]** | **98.6%** |
| Sex | -0.036 | [-0.090, 0.011] | 77.7% |
| Scanning protocol | 0 | [-0.061, 0.086] | 46.3% |
| Scanner | 0 | [-0.050, 0.055] | 58.2% |
| Medication | 0 | [0, 0] | 79.3% |
| Diagnosis x LLP_DMN_–posterior cingulate _DMN_ | 0.007 | [-0.004, 0.015] | 82.8% |
| Diagnosis x LLP_DMN_–medial temporal _DMN_ | 0 | [-0.012, 0] | 43.2% |
| **Diagnosis x LLP_DMN_–RLP_DMN_** | **0.012** | **[0.0001, 0.025]** | **76.1%** |
| Diagnosis x LLP_DMN_–L inferior temporal _DMN_ | 0 | [-0.011, 0] | 39.9% |
| Diagnosis x LLP_DMN_–R inferior temporal _DMN_ | 0 | [-0.013, 0.003] | 49.7% |
| Diagnosis x LLP_DMN_–mid thalamus _DMN_ | 0 | [-0.007, 0.014] | 51.8% |
| Diagnosis x LLP_DMN_–L posterior cerebellar _DMN_ | 0 | [-0.010, 0] | 29.9% |
| Diagnosis x LLP_DMN_–R posterior cerebellar _DMN_ | 0 | [-0.010, 0] | 32.9% |
| Diagnosis x LLP_DMN_–dorsal anterior cingulate _SN_ | 0 | [0, 0.016] | 28.9% |
| Diagnosis x LLP_DMN_–LaPFC_SN_ | 0 | [0, 0.018] | 35.7% |
| Diagnosis x LLP_DMN_–RaPFC_SN_ | 0 | [0, 0.013] | 34.2% |
| Diagnosis x LLP_DMN_–LLP_SN_ | 0.004 | [-0.002, 0.011] | 76.9% |
| Diagnosis x LLP_DMN_–RLP_SN_ | 0.001 | [-0.007, 0.006] | 46.1% |

Supplementary Table 3. Results from LASSO regression with bootstrap analysis identifying reliable predictors of lifetime tobacco use. Beta, average estimated regression coefficient across bootstrapping LASSO regression models; CI, confidence interval. Bolded parameters had statistically significant nonzero beta.

**Supplementary Table 4**

|  | Population | Estimate | SE | t-value | p-value |
| --- | --- | --- | --- | --- | --- |
| LLP_DMN_ – L hotspot | Ever-use | -2.764e-04 | 3.209e-04 | -0.861 | 0.3906 |
| LLP_DMN_ – R hotspot | Ever-use | -8.177e-04 | 4.613e-04 | -1.773 | 0.07863 |

Supplementary Table 4. Results from post-hoc linear regression investigating the relationship between diagnosis-by-daily cigarette use interaction and LLP_DMN_ – hotspot connectivity. SE, standard error.

**Supplementary Table 5**

|  | r | DoF | t-value | p-value |
| --- | --- | --- | --- | --- |
| Among participants with ever-use | | | | |
| All with ever-use (n=137) | 0.2032 | 135 | 2.411 | 0.01725* |
| Psychosis with ever-use (n=106) | 0.1624 | 104 | 1.6787 | 0.09622 |
| Control with ever-use (n=31) | 0.3349 | 29 | 1.9139 | 0.06555 |
| Among participants with current-use | | | | |
| All with current-use (n=92) | 0.2232 | 90 | 2.1727 | 0.03243* |
| Psychosis with current-use (n=75) | 0.1250 | 73 | 1.0766 | 0.2852 |
| Control with current-use (n=17) | 0.6639 | 15 | 3.4386 | 0.003657** |

Supplementary Table 5. Results from post-hoc Pearson’s correlation investigating the relationship between daily cigarette use and LLP_DMN_ – right posterior insula connectivity. DoF, degree of freedom; r, Pearson’s coefficient. *: 0.01<p<0.05, **: 0.001<p<0.01, ***: p<0.001.
